# Effectiveness of a digital health application (*levidex*) on quality of life in people with multiple sclerosis: A pragmatic, randomized controlled trial (LAMONT)

**DOI:** 10.64898/2026.03.12.26348037

**Authors:** Björn Meyer, Gereon Nelles, Linda Betz, Arnfin Bergmann, Kamila Jauch-Chara, Nicole Krause, Karin Riemann, Barbara von Glasenapp, Christoph Heesen

## Abstract

**Background:** People with multiple sclerosis (pwMS) often experience impaired quality of life (QoL) despite receiving standard care. Digital therapeutics (DTx) may offer support, but prior trials yielded mixed results, possibly due to active controls and high baseline QoL. We therefore evaluated a DTx (*levidex*) as an adjunct to treatment as usual (TAU) in pwMS with impaired QoL.

**Methods:** In this pragmatic, online randomised controlled trial (LAMONT; NCT06090305), *n* = 470 pwMS with a score ≥2 on the Hamburg Quality of Life Questionnaire in Multiple Sclerosis (HAQUAMS) were randomised to *levidex* + TAU or TAU alone. The primary endpoint was HAQUAMS total score at 6 months, analysed by intention-to-treat ANCOVA.

**Results:** Compared with TAU, *levidex* + TAU improved MS-specific QoL at 6 months (baseline-adjusted mean difference −0.10; 95% CI −0.18 to −0.03; p = 0.008; Cohen’s *d* = 0.26). Clinically relevant HAQUAMS improvement (≥0.22) occurred more often with *levidex* (39.5% vs 27.8%; number needed to treat = 9). Benefits also emerged for depressive symptoms and social/work functioning but not for anxiety. No serious adverse events occurred and user satisfaction was high.

**Conclusions:** In pwMS with impaired QoL, adding the scalable DTx *levidex* to TAU yields meaningful improvements in QoL and functioning.

## Introduction

Multiple sclerosis (MS) is a chronic, inflammatory, neurodegenerative disease affecting an estimated 2.8 million people globally.^1^ Beyond sensory and motor symptoms, fatigue, cognitive difficulties, and emotional distress profoundly impact health-related quality of life (QoL).^2^ While disease-modifying drugs (DMDs) can reduce inflammation and relapses, they do not directly address these psychosocial and emotional aspects, which are sometimes termed “invisible symptoms”.^3^ Accordingly, guidelines increasingly recommend holistic approaches, including non-pharmacological approaches like cognitive behavioural therapy (CBT) and health-behaviour interventions (e.g., physical activity, diet, stress management) to improve QoL.^4,5^

Access to psychological and behavioural support is limited for many people with MS (pwMS) due to therapist shortages, geographical barriers, and disability-related constraints like reduced mobility and fatigue.^6^ Digital therapeutics (DTx) may help by delivering evidence-based interventions remotely, although efficacy and safety need to be established for each program.^7^ Despite a growing DTx landscape, few interventions are designed to address the multi-domain needs of pwMS.

*levidex* is a DTx tailored for pwMS and is evaluated in this randomized controlled trial (RCT). The program’s development and initial pilot testing confirmed its relevance, trustworthiness, and perceived utility for inspiring healthy lifestyle changes, as reported by pwMS and clinicians.^8^ Two previous RCTs investigated the program’s efficacy, but with notable limitations. The first compared *levidex* to web-adapted informational material, finding significant but small improvements in QoL, sick leave, and daily living activities.^9^ However, the trial suffered from high dropout and, crucially, its active control group precluded comparison to standard treatment as usual (TAU). The second RCT compared *levidex* to a non-personalised active psychoeducational program (“dexilev”) in recently diagnosed pwMS and failed to meet its ambitious primary clinical endpoint (time to relapse or new lesion on magnetic resonance imaging), and no differential QoL effects emerged, possibly due to the sample’s only minor baseline QoL impairments.^10^

Both RCTs were limited by the inclusion of participants with high baseline QoL, creating potential ceiling effects. Furthermore, neither trial could determine the effect of *levidex* as an add-on to TAU, which reflects its intended real-world use. Despite the ambiguous QoL findings in the second trial, related quantitative and qualitative data suggest *levidex* users report greater health-behaviour changes (e.g., diet, stress management) than controls.^11,12^

Previous research, therefore, left two critical questions unanswered: 1) the efficacy of *levidex* as an adjunct to true TAU (instead of an active control), and 2) its impact on an MS-population with pre-existing QoL impairment, thus avoiding ceiling effects.

The current trial (the LAMONT trial) addresses these gaps. We aimed to evaluate whether adding *levidex* to TAU improves MS-specific health-related QoL at 6 months compared with TAU alone in a population screened for impaired QoL. We hypothesised that *levidex* + TAU would be superior to TAU alone for the primary QoL outcome and for secondary outcomes, including mood, anxiety, functioning, and health utilisation.

## Methods

### Trial Design

We conducted a randomised, parallel-group superiority trial (LAMONT; NCT06090305) comparing levidex + treatment as usual (TAU) versus TAU alone. The primary endpoint was MS-specific QoL at 6 months (Hamburg Quality of Life Questionnaire in Multiple Sclerosis, HAQUAMS)^13^. Secondary endpoints included depressive symptoms, anxiety, social/work functioning, MS-specific QoL (alternate instrument), and instrumental activities of daily living.

### Eligibility criteria

Eligible participants were ≥18 years with physician-confirmed MS (ICD-10-GM G35.x), impaired QoL (HAQUAMS total ≥2; a cut-off chosen to capture patients with at least moderate disability, approximating an Expanded Disability Status Scale [EDSS] score ≥3), specialist MS care within 3 months, and ability to use an online programme. Additional criteria were German language proficiency, internet access, and written informed consent. An exclusion criterion was severe dependence (Pflegegrad, §15 SGB XI) ≥3.

### Study Setting and Data Collection

The trial was conducted fully remotely (9 Nov 2023–6 Dec 2024). Participants were recruited via online campaigns/newsletters and physician referrals to a study website. Data were collected via LimeSurvey^14^ using range/validity checks; study procedures and data integrity were monitored regularly with routine backups.

### Interventions

#### Control Group (CG)

CG participants received TAU, reflecting routine outpatient care (e.g., primary/specialist care, DMDs/other medication, psychotherapy, or no active treatment). All CG particpants also received a brief digital information sheet with publicly available MS resources (e.g., German MS Society).

#### Intervention Group (IG): levidex

Following randomization, IG participants in the IG received access to *levidex* adjunctively to TAU. This is a web-based DTx for pwMS, developed, owned, and operated by GAIA, a small-to-medium enterprise specializing in DTx. As described previously, it was developed in collaboration with a multidisciplinary team of neurologists, clinical psychologists, health scientists, and nutritionists.^8^ The intervention is delivered via standard internet browsers on common devices (computer, tablet, smartphone) and is implemented on GAIA’s proprietary broca® platform, which uses rule-based algorithms to generate personalised “simulated dialogues.” Users progress through modules by selecting predefined responses; subsequent content is algorithmically tailored to user characteristics and preferences. The platform’s dialogue-based CBT approach has been shown to be effective in previous RCTs of other DTx, including for MS-related fatigue and depression.^15,16^

*levidex* comprises 16 sequential modules (“conversations”; ∼30–45 min each) covering: (1) MS self-management/education, (2) psychological strategies for emotional well-being, (3) dietary approaches, and (4) physical activity and sleep. Modules include in-program exercises (e.g. planning, mindfulness and relaxation or imagery-based audio exercises) and between-session activities (e.g. activity plans, preparing specific foods), supported by optional message reminders and staged module release to encourage task completion and reflection.

The program is primarily CBT-based and integrates common behaviour change techniques (BCTs), such as cognitive restructuring, behavioural activation, goal-setting, action planning, self-monitoring, provision of information about health consequences, and prompts to support health-promoting behaviours.^17^ In addition, *levidex* incorporates mindfulness- and acceptance-based components, mental imagery exercises, and elements of motivational interviewing, and is aligned with the concept of patient empowerment.^18^ Overall, *levidex* was designed with the aim of supporting pwMS in optimising health-related behaviours (e.g. physical activity, dietary behaviours, stress management and sleep) and improving health-related QoL.

### Outcomes

Outcome data were collected at 3 months (T1) and 6 months (T2) after randomisation, with the 6-month assessment prespecified as the primary evaluation time point to meet regulatory requirements for permanent inclusion in the German Digital Health Applications (DiGA) directory. The 3-month assessment was included to explore early effects. After completion of the T2 assessment, participants in the CG were offered access to *levidex*.

#### Primary outcome

The primary endpoint was MS-specific health-related QoL at 6 months (HAQUAMS total score).^13^

#### Secondary outcomes

Secondary endpoints included depressive symptoms (Patient Health Questionnaire-9, PHQ-9)^19^, social and work-related functioning (Work and Social Adjustment Scale, WSAS)^20^, MS-specific health-related quality of life (global index score of the Multiple Sclerosis International Quality of Life questionnaire, MusiQoL)^21^, anxiety symptoms (Generalized Anxiety Disorder Scale-7, GAD-7)^22^, and instrumental activities of daily living (Frenchay Activities Index, FAI).^23^

#### Exploratory outcomes

Exploratory endpoints included HAQUAMS subscales, overall DMD intake in the previous 3 months, and DMD intake classified according to efficacy (categories 1–3 per current German guideline). Additional exploratory endpoints were health care utilisation indicators (number of days on sick leave/sick pay and the number of days in inpatient treatment during the previous 3 months), and user satisfaction (assessed by the Net Promoter Score [NPS]^24^ and the ZUF-8 patient satisfaction questionnaire^25^).

### Sample Size

As pre-specified in the clinical investigation plan and trial registration, the trial aimed to include 470 pwMS, randomised in a 1:1 allocation ratio to receive *levidex* plus TAU or TAU alone. Reanalysis of data from a previous *levidex* RCT^9^ restricted to patients with impaired QoL (HAQUAMS ≥2) yielded an effect size of Cohen’s *d* = 0.28 for the HAQUAMS total score; assuming a slightly larger expected effect of *d* = 0.30, a total of 352 participants (176 per group) would provide 80% power at α = 0.05. Allowing for 25% attrition, we therefore aimed to recruit *N* = 470 participants.

### Randomisation

Participants were allocated 1:1 to the IG or the CG using simple randomisation via an external, web-based computerized tool. The allocation sequence was generated using computer-produced random numbers and was concealed from study staff; group assignment was released only after completion of baseline assessments. No blocking or stratification factors were used.

### Blinding

Due to the nature of the web-based intervention, neither participants nor investigators were blinded to group allocation. All outcomes were assessed via self-administered online questionnaires.

### Statistical methods

Details of the statistical analyses were pre-specified in the clinical investigation plan. The primary analysis followed the intention-to-treat (ITT) principle. The primary endpoint (HAQUAMS at 6 months) was analysed using ANCOVA with group as factor and baseline HAQUAMS as covariate; adjusted mean differences (95% CI) and p values are reported. Secondary continuous outcomes were analysed analogously; between-group effect sizes (Cohen’s *d*) were calculated from baseline-adjusted mean differences. Intermediate treatment effects at 3 months were examined using the same ANCOVA framework. Exploratory categorical outcomes were analysed using χ² tests.

Missing outcomes were handled under a missing-at-random assumption using bootstrapped maximum-likelihood multiple imputation (R packages *bootImpute* and *mice*),^26^ with imputation models including the respective baseline outcome, treatment group, and relevant sociodemographic and clinical variables (age, sex, MS type, concomitant psychotherapy, antidepressant use). A conservative reference-based sensitivity analysis (jump-to-reference imputation) was additionally performed for primary and secondary outcomes.^27^ Analyses were conducted using R (version 4.4.1)^28^, and two-sided *p* values <0.05 were considered statistically significant. To control for multiplicity across secondary outcomes, a hierarchical gatekeeping strategy was applied; if a test in the pre-specified sequence (PHQ-9, WSAS, MusiQoL, GAD-7, FAI) was not significant, subsequent endpoints were considered exploratory.

## Results

### Participants and follow-up

Participants were enrolled between November 2023 and June 2024 and followed for 6 months, with the last patient last visit on 6 December 2024. Numbers for screening, allocation, follow-up and analysis are shown in Figure 1. All 470 randomised participants were analysed in their originally assigned groups according to the ITT principle. Baseline characteristics are summarised in Table 1 and indicate broadly comparable groups.

**Figure 1:**
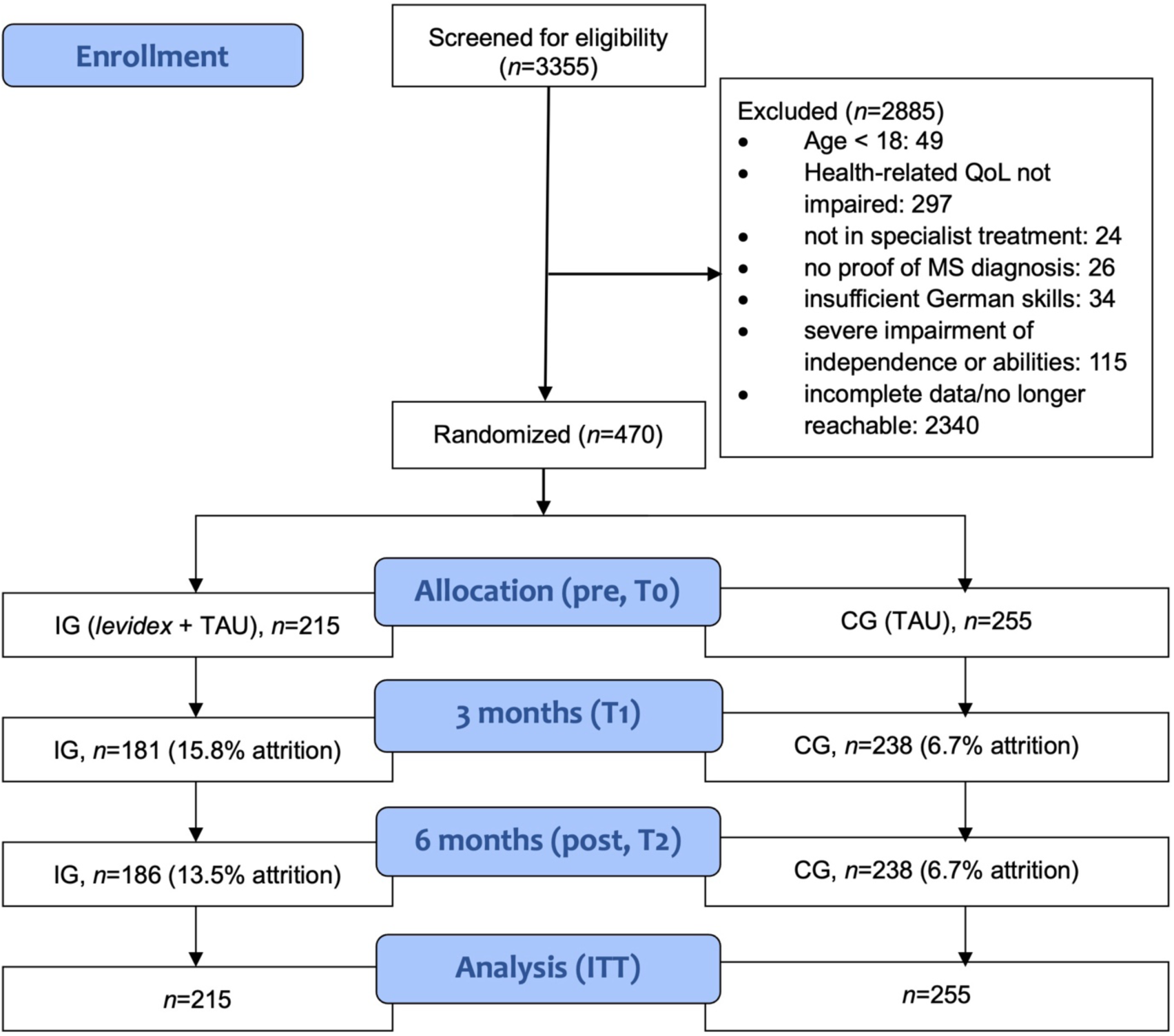
Participant flow

**Table 1:**
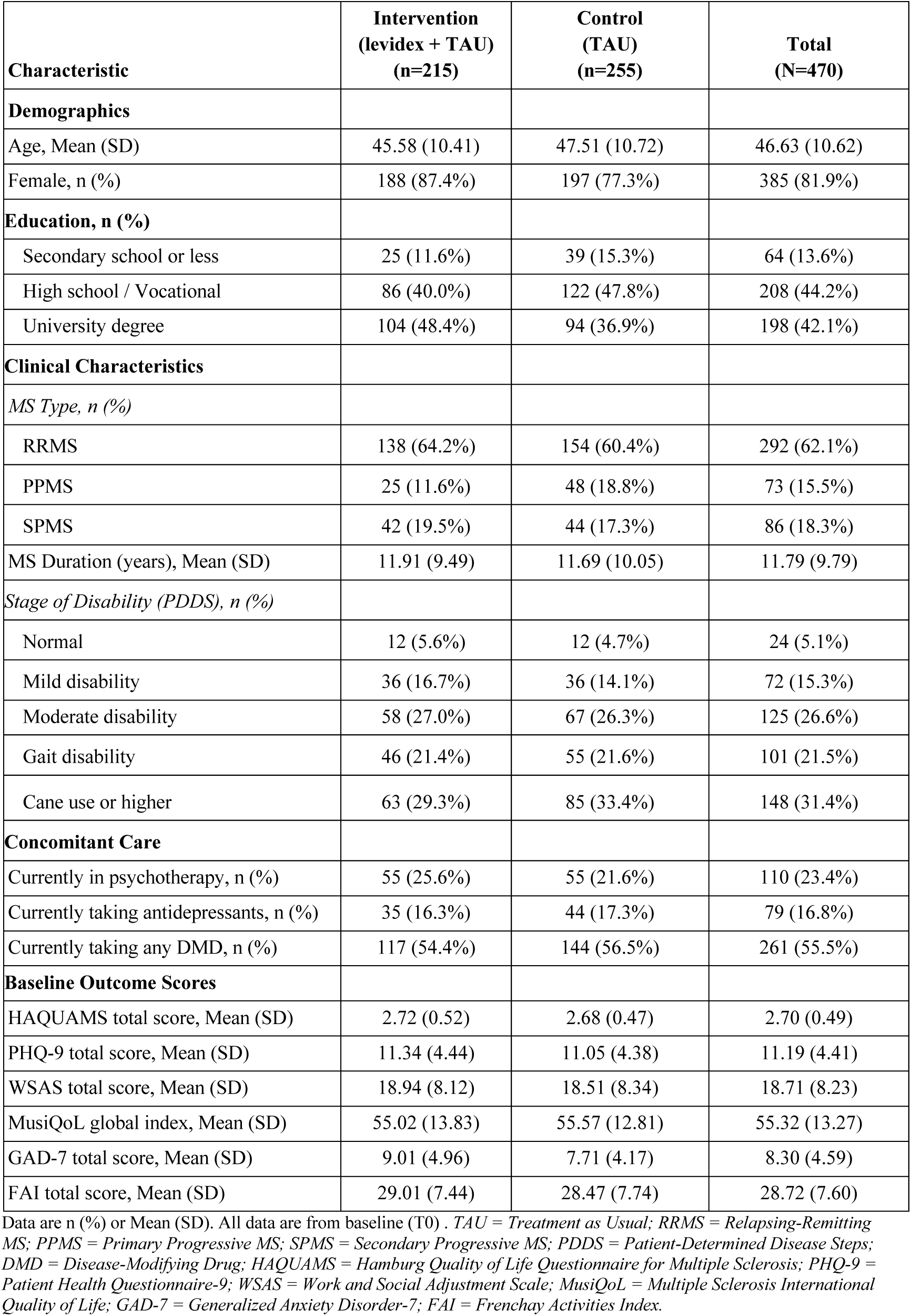
Baseline Sociodemographic and Clinical Characteristics (N=470)

### Primary outcome

In the ITT analysis, participants in the IG showed better MS-specific health-related QoL at 6 months than those in the CG (Table 2). The baseline-adjusted between-group difference on the HAQUAMS total score at T2 was −0.10 points (95% CI −0.18 to −0.03; *p* = 0.008; Cohen’s *d* = 0.26), with a similar effect in the sensitivity analysis (Supplementary Table 2). A higher proportion of pwMS in the IG achieved at least a minimal clinically important difference (MCID) of ≥0.22 points on the HAQUAMS than in the CG (39.5% vs 27.8%; OR 1.69, 95% CI 1.15–2.50; *p* = 0.007; NNT = 9). Improvements were already apparent at 3 months (Supplementary Table 1).

**Table 2:**
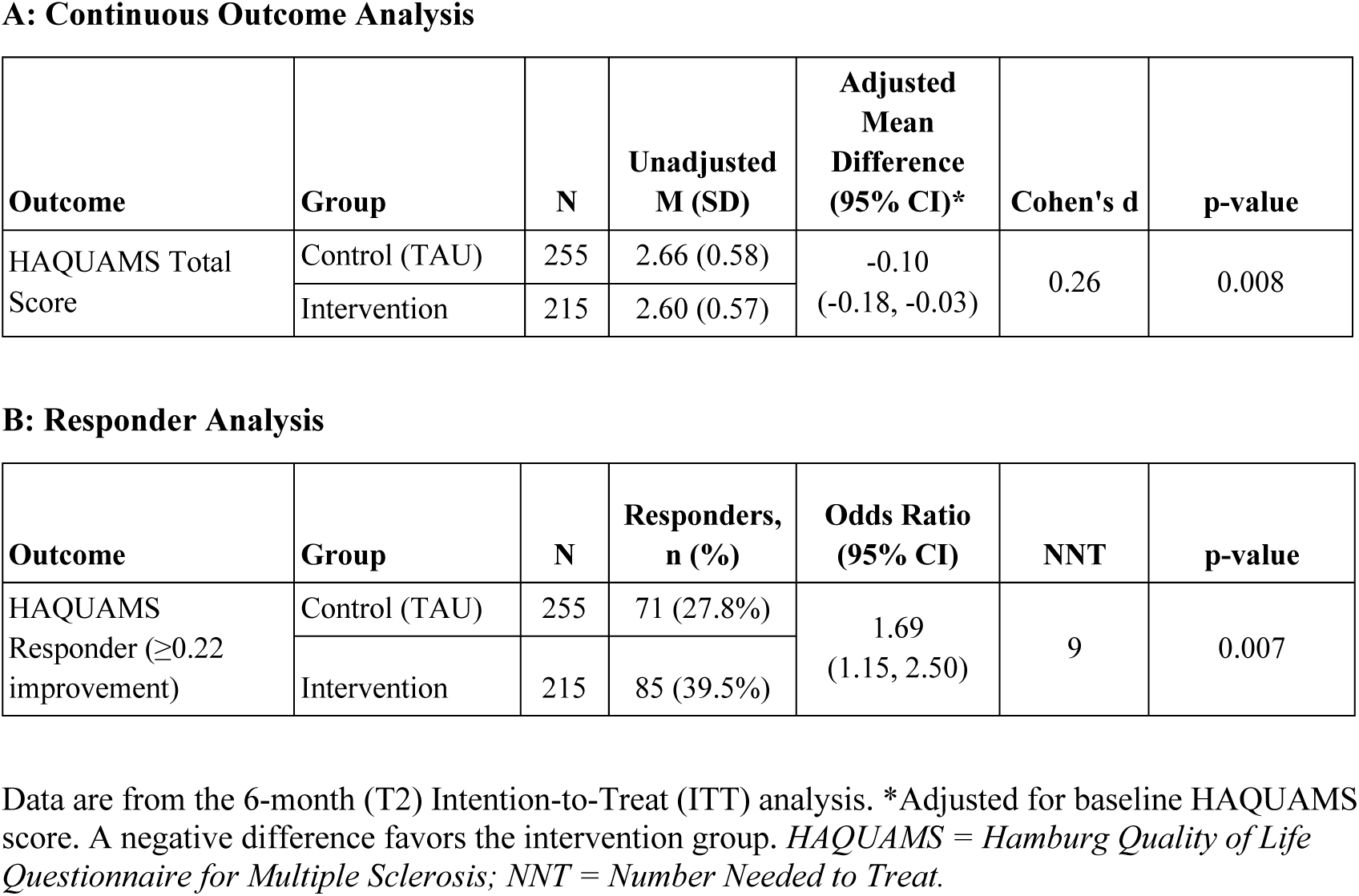
Primary Outcome (MS-Specific QoL, HAQUAMS) at 6 Months (ITT Analysis)

### Secondary outcomes

At 6 months (Table 3), *levidex* plus TAU yielded greater improvements than TAU alone in depressive symptoms (PHQ-9; baseline-adjusted mean difference −0.8 points, 95% CI −1.4 to −0.1; *p* = 0.025; *d* = 0.21), social and work functioning (WSAS; −1.8 points, 95% CI −2.9 to −0.6; *p* = 0.003; *d* = 0.30), and MS-specific health-related QoL assessed with the MusiQoL global index (2.1 points, 95% CI 0.3 to 3.9; *p* = 0.020; *d* = 0.23). Anxiety symptoms were not significantly different (GAD-7; baseline-adjusted mean difference −0.5 points, 95% CI −1.2 to 0.2; *p* = 0.205; *d* = 0.13). In line with the gatekeeping strategy, subsequent endpoints were treated as exploratory; for these outcomes, including instrumental activities of daily living (FAI; 0.8 points, 95% CI −0.1 to 1.8; *p* = 0.097; *d* = 0.17), no consistent intervention effects were detected. Responder analyses for secondary outcomes are available in Supplementary Table 3; results for exploratory HAQUAMS subscales and health utilization metrics in Supplementary Table 4 and Supplementary Table 5.

**Table 3:** Secondary Outcomes at 6 Months (ITT Analysis)

| **Outcome** | **Adjusted Mean Difference**<br>**(95% CI)\*** | ***p*-value** | **Cohen's *d*** |
| --- | --- | --- | --- |
| Depressive symptoms (PHQ-9) | -0.8 (-1.4, -0.1) | 0.025 | 0.21 |
| Social & work functioning (WSAS) | -1.8 (-2.9, -0.6) | 0.003 | 0.30 |
| MS-specific QoL (MusiQoL) | 2.1 (0.3, 3.9) | 0.02 | 0.23 |
| Anxiety symptoms (GAD-7) | -0.5 (-1.2, 0.2) | 0.205 | 0.13 |
| Instrumental activities (FAI)† | 0.8 (-0.1, 1.8) | 0.097 | 0.17 |

*Data are from the 6-month (T2) Intention-to-Treat (ITT) analysis. All differences are adjusted for their respective baseline scores. \*For PHQ-9, WSAS, and GAD-7, a negative difference favors the intervention. For MusiQoL and FAI, a positive difference favors the intervention.*
†Considered exploratory per the pre-specified gatekeeping strategy, as the analysis for anxiety symptoms (GAD-7) was not statistically significant. *PHQ-9* = *Patient Health Questionnaire-9*; *WSAS* = *Work and Social Adjustment Scale*; *MusiQoL* = *Multiple Sclerosis International Quality of Life*; *GAD-7* = *Generalized Anxiety Disorder-7*; *FAI* = *Frenchay Activities Index*.

### Intervention Adherence

All participants randomised to IG registered to use *levidex*. Over 6-month, participants completed a mean of 10.0 (*SD* 5.0) of the 16 available modules and had 25.1 (*SD* 23.7) days of active use.

### User Satisfaction

User satisfaction was assessed in the IG at 3 and 6 months. The NPS was 35.6 at T1 and 31.5 at T2, indicating very high user satisfaction (on a scale of -100 to 100). This was supported by the ZUF-8 patient satisfaction questionnaire, where the mean item score was 3.2 at both T1 (SD=4.1) and T2 (SD=5.0), reflecting high satisfaction (on a scale of 1-4).

### Ancillary analyses

Exploratory subgroup analyses of the primary outcome (HAQUAMS total score at 6 months) were conducted by sex, age, MS type, psychotherapy status, antidepressant use and DMD use at baseline. Overall, effects of *levidex* were in the same direction across most subgroups, with statistically significant improvements observed in women but not men and in participants taking antidepressants or not receiving DMD treatment at baseline, whereas effects were smaller and non-significant in those on DMDs or not using antidepressants (Supplementary Table 6 for full subgroup data). Exploratory analyses of DMD use at 3 and 6 months are presented in Supplementary Table 7 and did not show consistent differences between groups.

### Harms

No adverse events, adverse device effects, device deficiencies or serious adverse events were observed during the trial.

## Discussion

This pragmatic, online RCT found that adding the DTx *levidex* to TAU improved QoL in adults with MS. *levidex* produced small but statistically significant and clinically relevant MS-specific QoL improvements at 3 and 6 months compared with TAU alone (NNT=9 for HAQUAMS improvement). Additional benefits were observed for depressive symptoms, social/work functioning, and MS-specific QoL on MusiQoL; no significant effects were found for anxiety or instrumental activities of daily living. The intervention was well tolerated, with high user satisfaction (NPS at 6 months = 31.5). These findings suggest that this automated program can yield meaningful patient-reported gains as an adjunct to routine care.

A key implication is that this automated, multi-domain program can achieve clinically relevant improvements despite the challenges of sustained behaviour change in MS. The small-to-moderate effect sizes are comparable to those from more resource-intensive psychosocial interventions, while scalability and ease of implementation are substantially higher.^29,30^ Future work should examine whether enhanced tailoring and optional stepped support (including human guidance for those with greater needs) can increase benefit while preserving digital-first efficiency.

An earlier *levidex* trial in a recently diagnosed MS cohort with only mild QoL impairment, (baseline HAQUAMS about 1.7-1.8) and therefore limited room for improvement, focused on inflammatory activity and showed no advantage over an active control and no QoL difference.^10^ In contrast, another trial that recruited pwMS with more pronounced QoL impairment (baseline HAQUAMS about 2.5) reported a small but significant effect on QoL.^9^ The present study likewise enrolled pwMS with impaired QoL (baseline HAQUAMS 2.6) and found a comparable effect size. Although this mean difference is below the HAQUAMS MCID of 0.22 at the group level, almost 40% of pwMS in the *levidex* group achieved at least an MCID-level improvement compared with 28% under TAU (NNT = 9). Taken together, the trials suggest that *levidex* can serve as a useful adjunctive tool for pwMS experiencing reduced QoL, including when used alongside pharmacological treatment.

In Germany, the program has been permanently included in the national DiGA directory based on this pivotal trial, making it prescribable and reimbursable.^31^ However, reimbursement alone may not ensure uptake, and implementation efforts may be needed. This context underscores the relevance of these findings, as many pwMS report impaired QoL, residual symptoms, and difficulty maintaining healthy behaviours despite DMDs.^3^ This self-guided, low-threshold program may help to narrow the gap between pharmacological treatment and the daily demands of living with MS. *levidex* could serve as an early, scalable adjunctive option, with more intensive interventions reserved for non-responders.

The mechanism of *levidex* remains an important question. The multi-target approach (mood, coping, physical activity, diet, stress) suggests combined pathways rather than a single “active ingredient.” With self-reported outcomes and no objective behavioural indicators, mediation by behaviour change cannot be determined; future work should incorporate behavioural measures and mediation analyses.

Despite notable strengths, such as low attrition, recruitment of patients with impaired QoL, and the use of a true TAU comparison, several limitations must be acknowledged. First, recruitment may have attracted motivated, digitally literate pwMS. While this limits generalisability to the entire MS population, it also reflects the target population for whom a self-guided digital intervention like *levidex* is intended and most appropriate. Second, all outcomes were self-reported, lacking objective verification (e.g., accelerometry). While this introduces a potential for reporting bias, patient perception is arguably the most valid and direct measure for subjective constructs like QoL and depressive symptoms. Moreover, recent evidence suggests that self-report instruments in behavioural intervention trials do not necessarily overestimate effects and may even be more conservative than clinician assessments.^32^ Consistent with this, a process evaluation of a *levidex* trial showed that meaningful behaviour changes were often reported qualitatively but not fully reflected in standard questionnaires, indicating that commonly used outcome measures may lack sensitivity for lifestyle-related changes.^12^ Third, the 6-month follow-up limits conclusions on sustainability or effects on harder clinical outcomes. Fourth, the unblinded, TAU-alone control design cannot separate specific intervention effects from non-specific factors like expectancy or study participation; however, this design is aligned with methodological recommendations and was necessary to assess real-world additive benefit.^33^ Finally, the German-speaking cohort requires replication in other settings, and subgroup analyses were not powered for definitive conclusions.

In conclusion, adding *levidex* to TAU yielded small but meaningful improvements in MS-specific QoL, mood, and functioning in adults with MS and impaired QoL. While effects are modest, scalability and low demands on clinical resources suggest population-level value. Future research should refine implementation in stepped-care models, test longer-term and objective outcomes, and evaluate whether enhanced tailoring or added support improves effectiveness.

## Acknowledgements

The authors wish to thank all individuals with MS who participated in the LAMONT trial. We also extend our gratitude to the neurologists and treating physicians who supported recruitment by referring their patients to this fully remote study.

## Protocol

The design and statistical analyses were registered as part of the trial registration (ClinicalTrials.gov: NCT06090305) before data analysis.

## Monitoring

This fully remote trial did not involve external independent data monitoring. Data quality and study procedures were monitored internally by the sponsor (Gaia AG), including regular data integrity checks and monitoring of participant follow-up, as described in the Methods.

## Author contributions

KJC is the principal investigator and led the conception, planning, and conduct of the study. BM wrote the first draft of the manuscript and contributed to trial design and interpretation. LB contributed to data analysis, trial design, data acquisition, and interpretation. GN and CH contributed to the study’s conception and provided critical intellectual revisions. All authors revised the manuscript for important intellectual content and approved the final version for publication.

## Data availability statement

The data that support the findings of this study are available from the sponsor (Gaia AG) upon reasonable request and subject to a data-sharing agreement.

## Ethical considerations

The study protocol, clinical investigation plan, and all participant-facing materials, including the digital informed consent procedure, were approved by the responsible independent Ethics Committee of the Hamburg Chamber of Physicians (Ethik-Kommission der Ärztekammer Hamburg; reference number 2023-101078-BO-ff). The trial was conducted in accordance with the Declaration of Helsinki and all applicable German regulations.

## Consent to participate

Informed consent was obtained digitally via the secure study website. Interested individuals were presented with detailed, comprehensive study information approved by the ethics committee. Before any study procedures or randomization, participants were required to confirm their understanding and provide written informed consent electronically (by checking mandatory confirmation boxes and providing a digital signature). Participants were informed that they could withdraw their consent at any time without disadvantage. Participants were compensated with a €10 gift voucher for each follow-up assessment they completed. As stated in the informed consent, participants in the control group were offered access to the *levidex* program free of charge after completion of the 6-month primary endpoint assessment.

## Supplemental material

Supplemental material for this article is available online.

