## Supplementary material for "Effectiveness of a digital health application (*levidex*) on quality of life in people with multiple sclerosis: A pragmatic, randomized controlled trial (LAMONT)": Supplmentary Materials

**Supplementary Table 1:** Primary and Secondary Outcomes at 3 Months (T1) (ITT Analysis)

| **Outcome** | **Adjusted Mean Difference (95% CI)*** | ***p*-value** | **Cohen's *d*** |
|---|---|---|---|
| MS-Specific QoL (HAQUAMS) | -0.11 (-0.18, -0.05) | < .001 | 0.32 |
| Depressive symptoms (PHQ-9) | -0.2 (-0.9, 0.5) | 0.63 | 0.05 |
| Social & work functioning (WSAS) | -1.7 (-2.7, -0.6) | 0.002 | 0.3 |
| MS-specific QoL (MusiQoL) | 1.5 (-0.1, 3.1) | 0.071 | 0.18 |
| Anxiety symptoms (GAD-7) | 0.2 (-0.5, 0.9) | 0.537 | -0.06 |
| Instrumental activities (FAI) | 0.4 (-0.5, 1.3) | 0.418 | 0.08 |

*Data are from the 3-month (T1) Intention-to-Treat (ITT) analysis. All differences are adjusted for their respective baseline scores. *For HAQUAMS, PHQ-9, WSAS, and GAD-7, a negative difference favors the intervention. For MusiQoL and FAI, a positive difference favors the intervention. HAQUAMS = Hamburg Quality of Life Questionnaire for Multiple Sclerosis; PHQ-9 = Patient Health Questionnaire-9; WSAS = Work and Social Adjustment Scale; MusiQoL = Multiple Sclerosis International Quality of Life; GAD-7 = Generalized Anxiety Disorder-7; FAI = Frenchay Activities Index.*

**Supplementary Table 2:** Conservative Sensitivity Analyses (Jump-to-Reference Imputation) for Primary and Secondary Outcomes at 6 Months (T2)

| Outcome | Adjusted Mean Difference (95% CI)* | p-value | Cohen's d |
|---|---|---|---|
| **Primary Outcome** | | | |
| MS-Specific QoL (HAQUAMS) | -0.09 (-0.16, -0.03) | 0.003 | 0.24 |
| **Secondary Outcomes** | | | |
| Depressive symptoms (PHQ-9) | -0.6 (-1.2, -0.1) | 0.027 | 0.18 |
| Social & work functioning (WSAS) | -1.4 (-2.3, -0.5) | 0.002 | 0.25 |
| MS-specific QoL (MusiQoL) | 1.9 (0.5, 3.3) | 0.008 | 0.21 |
| Anxiety symptoms (GAD-7) | -0.5 (-1.0, 0.1) | 0.088 | 0.14 |
| Instrumental activities (FAI) | 0.6 (-0.2, 1.3) | 0.163 | 0.12 |

*Data are from the 6-month (T2) Jump-to-Reference (J2R) sensitivity analysis. All differences are adjusted for their respective baseline scores. *For HAQUAMS, PHQ-9, WSAS, and GAD-7, a negative difference favors the intervention. For MusiQoL and FAI, a positive difference favors the intervention. HAQUAMS = Hamburg Quality of Life Questionnaire for Multiple Sclerosis; PHQ-9 = Patient Health Questionnaire-9; WSAS = Work and Social Adjustment Scale; MusiQoL = Multiple Sclerosis International Quality of Life; GAD-7 = Generalized Anxiety Disorder-7; FAI = Frenchay Activities Index.*

**Supplementary Table 3:** Responder Analyses for Secondary Outcomes at 6 Months (T2) (ITT Analysis)

| Outcome | Responder Definition (MCID) | Group | N | Responders, n (%) | Odds Ratio (95% CI) | p-value |
|---|---|---|---|---|---|---|
| Depressive symptoms (PHQ-9) | ≥5 point improvement | Control (TAU) | 255 | 52 (20.4%) | 1.12 (0.72, 1.75) | 0.61 |
| | | Intervention | 215 | 48 (22.3%) | | |
| Social & work functioning (WSAS) | ≥8 point improvement | Control (TAU) | 255 | 16 (6.3%) | 2.80 (1.50, 5.24) | < .001 |
| | | Intervention | 215 | 34 (15.8%) | | |
| MS-specific QoL (MusiQoL) | ≥7.375 point improvement (0.5 SD) | Control (TAU) | 255 | 56 (22.0%) | 1.64 (1.09, 2.48) | 0.018 |
| | | Intervention | 215 | 68 (31.6%) | | |

Data are from the 6-month (T2) Intention-to-Treat (ITT) analysis. *MCID = Minimal Clinically Important Difference; SD = Standard Deviation. PHQ-9 = Patient Health Questionnaire-9; WSAS = Work and Social Adjustment Scale; MusiQoL = Multiple Sclerosis International Quality of Life.*

**Supplementary Table 4:** Exploratory Outcomes (HAQUAMS Subscales) at 6 Months (T2) (ITT Analysis)

| Outcome (HAQUAMS Subscale) | Adjusted Mean Difference (95% CI)* | p-value | Cohen's d |
|---|---|---|---|
| Cognition | -0.10 (-0.23, 0.03) | 0.121 | 0.14 |
| Fatigue | -0.12 (-0.26, 0.02) | 0.098 | 0.17 |
| Mobility (lower limb) | -0.14 (-0.26, -0.01) | 0.028 | 0.22 |
| Mobility (upper limb) | -0.03 (-0.12, 0.05) | 0.46 | 0.07 |
| Communication | -0.11 (-0.22, 0.00) | 0.052 | 0.19 |
| Mood | -0.08 (-0.20, 0.03) | 0.166 | 0.14 |

*Data are from the 6-month (T2) Intention-to-Treat (ITT) analysis. All differences are adjusted for their respective baseline scores. *A negative difference favors the intervention group. HAQUAMS = Hamburg Quality of Life Questionnaire for Multiple Sclerosis.*

**Supplementary Table 5:** Exploratory Outcome: Health Utilization at 3 and 6 Months (Complete Case Analysis)

| **Outcome (previous 3 mos.)** | **Time** | **Group** | **N** | **Mean (SD)** | **W** | **p-value** |
|---|---|---|---|---|---|---|
| Days on sick leave | T1 (3 mos) | Control (TAU) | 238 | 15.4 (28.7) | 19500 | 0.078 |
| | | Intervention | 181 | 18.8 (31.4) | | |
| | T2 (6 mos) | Control (TAU) | 237 | 13.3 (26.8) | 21080 | 0.411 |
| | | Intervention | 186 | 15.8 (28.9) | | |
| Days on sick pay | T1 (3 mos) | Control (TAU) | 238 | 7.2 (22.9) | 19722 | 0.019 |
| | | Intervention | 181 | 10.8 (27.4) | | |
| | T2 (6 mos) | Control (TAU) | 238 | 5.2 (19.3) | 22216 | 0.904 |
| | | Intervention | 186 | 7.2 (23.3) | | |
| Days in inpatient treatment | T1 (3 mos) | Control (TAU) | 238 | 1.5 (5.8) | 21356 | 0.797 |
| | | Intervention | 181 | 1.4 (5.4) | | |
| | T2 (6 mos) | Control (TAU) | 237 | 1.6 (6.8) | 21837 | 0.775 |
| | | Intervention | 186 | 1.8 (9.9) | | |

*Health utilization data (A) analyzed using Wilcoxon-Rank Sum Test.*

**Supplementary Table 6:** Subgroup Analyses for Primary Outcome (HAQUAMS Total Score) at 6 Months (T2) (ITT Analysis)

| **Subgroup** | **N** | **Adjusted Mean Difference (95% CI)*** | **p-value** |
|---|---|---|---|
| Overall Effect | 470 | -0.10 (-0.18, -0.03) | 0.008 |
| ***Age*** | | | |
| 18-65 years | 457 | -0.10 (-0.17, -0.02) | 0.012 |
| > 65 years | 13 | Not calculated (unstable analysis) | n/a |
| ***Sex*** | | | |
| Women | 385 | -0.11 (-0.19, -0.03) | 0.007 |
| Men | 85 | -0.02 (-0.17, 0.13) | 0.83 |
| ***MS-Type*** | | | |
| RRMS | 292 | -0.09 (-0.19, 0.00) | 0.057 |
| PPMS | 73 | -0.11 (-0.28, 0.05) | 0.168 |
| SPMS | 86 | -0.09 (-0.24, 0.06) | 0.219 |
| ***Psychotherapy Status*** | | | |
| In psychotherapy | 110 | -0.10 (-0.27, 0.07) | 0.238 |
| Not in psychotherapy | 360 | -0.10 (-0.18, -0.02) | 0.016 |
| ***Medication (Antidepressants)*** | | | |
| On antidepressants | 79 | -0.25 (-0.41, -0.08) | 0.004 |
| Not on antidepressants | 391 | -0.07 (-0.15, 0.01) | 0.093 |
| ***DMD Use*** | | | |
| On any DMD | 261 | -0.06 (-0.15, 0.04) | 0.233 |
| Not on DMD | 209 | -0.16 (-0.27, -0.05) | 0.006 |

Data are from the 6-month (T2) Intention-to-Treat (ITT) analysis. All differences are adjusted for baseline HAQUAMS scores. **A negative difference favors the intervention group (levidex + TAU). HAQUAMS = Hamburg Quality of Life Questionnaire for Multiple Sclerosis; RRMS = Relapsing-Remitting MS; PPMS = Primary Progressive MS; SPMS = Secondary Progressive MS; DMD = Disease-Modifying Drug.*

***Note:*** *The subgroup of participants >65 years (n=13) was too small for a stable ANCOVA*

***Supplementary Table 6:*** *Exploratory Outcome: DMD Use at 3 and 6 Months (Complete Case Analysis)*

| **Outcome (previous 3 mos.)** | **Time** | **Group** | **N** | **Yes, n (%)** | **Odds Ratio (95% CI)** | **χ2** | **p-value** |
|---|---|---|---|---|---|---|---|
| DMDs (Overall) | T1 (3 mos) | Control (TAU) | 238 | 124 (52.1%) | 1.30 (0.88, 1.92) | 1.73 | 0.188 |
| | | Intervention | 181 | 106 (58.6%) | | | |
| | T2 (6 mos) | Control (TAU) | 238 | 123 (51.7%) | 1.21 (0.82, 1.78) | 0.96 | 0.328 |
| | | Intervention | 186 | 105 (56.5%) | | | |
| DMDs (Category 1)[a] | T1 (3 mos) | Control (TAU) | 238 | 42 (17.6%) | 1.54 (0.96, 2.48) | 3.25 | 0.071 |
| | | Intervention | 181 | 45 (24.9%) | | | |
| | T2 (6 mos) | Control (TAU) | 238 | 37 (15.5%) | 1.63 (1.00, 2.66) | 3.91 | 0.048 |
| | | Intervention | 186 | 43 (23.1%) | | | |
| DMDs (Category 2)[b] | T1 (3 mos) | Control (TAU) | 238 | 17 (7.1%) | 1.71 (0.87, 3.34) | 2.48 | 0.115 |
| | | Intervention | 181 | 21 (11.6%) | | | |
| | T2 (6 mos) | Control (TAU) | 238 | 20 (8.4%) | 1.17 (0.60, 2.28) | 0.21 | 0.649 |
| | | Intervention | 186 | 18 (9.7%) | | | |
| DMDs (Category 3)[c] | T1 (3 mos) | Control (TAU) | 238 | 65 (27.3%) | 0.78 (0.50, 1.22) | 1.18 | 0.278 |
| | | Intervention | 181 | 41 (22.7%) | | | |
| | T2 (6 mos) | Control (TAU) | 238 | 66 (27.7%) | 0.81 (0.52, 1.26) | 0.9 | 0.342 |
| | | Intervention | 186 | 44 (23.7%) | | | |

χ2-test was used for analysis. *DMD = Disease-Modifying Drug.*

[a]: Efficacy category 1 *(e.g., dimethyl fumarate, glatirameroids, interferon-beta, teriflunomide)*

[b]: Efficacy category 2 *(e.g., cladribine, fingolimod, ozanimod, ponesimod, siponimod)*

[c]: Efficacy category 3 *(e.g., alemtuzumab, natalizumab, ocrelizumab, ofatumumab, rituximab, ublituximab)*
